# Scalable depression monitoring with smartphone speech: a multimodal benchmark and topic analysis

**DOI:** 10.1101/2025.07.17.25331744

**Authors:** Daniel Emden, Maike Richter, Astrid Chevance, Ramona Leenings, Julian Herpertz, Lara Gutfleisch, Anna Fleuchaus, Rogério Blitz, Vincent L. Holstein, Janik Goltermann, Nils R. Winter, Jennifer Spanagel, Susanne Meinert, Tiana Borgers, Kira Flinkenflügel, Frederike Stein, Nina Alexander, Hamidreza Jamalabadi, Jonathan Repple, Christian Dobel, Elisabeth J. Leehr, Ronny Redlich, Ulrich Ebner-Priemer, Igor Nenadić, Tilo Kircher, Udo Dannlowski, Tim Hahn, Nils Opel

**Affiliations:** Department of Psychiatry and Psychotherapy, Jena University Hospital, Jena, Germany; Institute for Translational Psychiatry, University of Münster, Münster, Germany; Department of Psychiatry and Neuroscience, Campus Benjamin Franklin, Charité–Universitätsmedizin Berlin, Berlin, Germany; Université Paris Cité and Université Sorbonne Paris Nord, INSERM INRAE, Centre for Research in Epidemiology and Statistics, Paris, France; Centre d’Epidémiologie Clinique, AP-HP, Hôpital Hôtel Dieu, Paris, France; Department of Psychiatry, Harvard Medical School, Boston, MA, USA; Stanley Center for Psychiatric Research, Broad Institute of MIT and Harvard, Cambridge, MA, USA; Department of Psychiatry and Psychotherapy, Philipps-University of Marburg, Marburg, Germany; Department of Psychiatry, Psychosomatic Medicine and Psychotherapy, University Hospital Frankfurt, Frankfurt am Main, Germany; Cooperative Brain Imaging Center - CoBIC, Goethe University Frankfurt, Frankfurt am Main, Germany; Department of Otorhinolaryngology, Institute of Phoniatry and Pedaudiology, Jena University Hospital, Jena, Germany; Department of Psychology, Martin Luther University Halle-Wittenberg, Halle, Germany; Center for Intervention and Research on adaptive and maladaptive brain Circuits underlying mental health (C-I-R-C), Jena-Magdeburg-Halle, Germany; German Center for Mental Health (DZPG), Germany; Mental mHealth Lab, Institute of Sports and Sports Science, Karlsruhe Institute of Technology, Karlsruhe, Germany; Center for Mind, Brain and Behavior (CMBB), Philipps-University of Marburg, Marburg, Germany

**Keywords:** Major Depressive Disorder, Smartphone Speech, Digital Phenotyping, Natural Language Processing, Large Language Models, Sentence Embeddings, Topic Modelling

## Abstract

Objective, scalable biomarkers are needed for continuous monitoring of major depressive disorder (MDD). Smartphone-collected speech is promising, yet extracting clinically useful signals remains difficult. We analysed 3 151 weekly voice diaries from 284 German-speaking adults (128 MDD, 156 controls) and regressed Beck Depression Inventory (BDI) scores. Sentence embeddings from the open-source 8-billion-parameter Qwen3-8B model predicted scores with MAE = 4.45 and *R*^2^ = 0.35, explaining 16 more points of variance than the best traditional feature set (TF-IDF). Adding lexical–prosodic or TF-IDF features provided only marginal improvement (best MAE = 4.39). To interpret the embeddings we applied BERTopic and uncovered ten coherent themes; BDI scores peaked for “Persistent Low Mood” and “Pain Distress”, confirming clinical relevance. Large-language-model embeddings therefore capture the dominant signal of depression severity in everyday speech and, paired with interpretable topic analysis, offer a privacy-preserving, scalable route to digital mental-health phenotyping.

## 1 Introduction

Depression is a leading contributor to the global burden of disease, defined by its chronic and recurrent nature. Consequently, long-term observation is crucial for prevention, yet its clinical course is typically monitored through infrequent, clinic-based self-reports [1, 2]. This “snapshot” view often misses the day-to-day fluctuations that can signal relapse or treatment response. Digital phenotyping seeks to close that gap by continuously harvesting data from personal devices, turning everyday behaviour into continuous mental-health read-outs [3, 4]. Among the many candidate signals, spoken language is unique: it is produced spontaneously, carries rich semantic content, and embeds prosodic cues that mirror affect and cognition [5, 6]. Voice diaries recorded in naturalistic (ecologically valid) contexts therefore offer a non-invasive lens on mood that can scale to the population level.

Early speech-based studies for depression focused on handcrafted acoustic markers or word-count approaches. While informative, these shallow features capture only limited facets of meaning and often fail to generalise across speakers and conditions. The paradigm shift towards transformer-based language models [7] now provides embeddings from large, pre-trained language models that compress nuanced semantics into dense vectors. These methods are increasingly being applied to mental health analysis [8, 9]. Yet few works have systematically benchmarked such embeddings against traditional lexical and acoustic pipelines on naturalistic data.

Equally important is the challenge of interpretability. Black-box predictions risk limited uptake in clinical settings unless researchers can articulate why a model outputs a high severity score [10]. Topic modelling methods such as BERTopic [11], an algorithm that automatically analyzes text to identify underlying discussion topics, bridge this gap by clustering embeddings into coherent themes whose prevalence can be related back to symptoms. This dual focus on performance plus insight is critical if digital phenotyping tools are to inform therapeutic decision-making rather than merely flag anomalies [12].

While promising, research in speech-based depression assessment has historically faced key limitations. Much of the foundational work has focused on classifying individuals with major depression versus healthy controls, often using speech elicited in controlled, laboratory settings (e.g., reading tasks or structured interviews) [13, 14]. This approach, while crucial for establishing a signal, does not capture the day-to-day symptom variability within a clinical population. More recent digital phenotyping studies have begun to leverage data from personal devices, but often remain cross-sectional or have yet to robustly model symptom severity from speech collected naturalistically over time [15, 16]. The present study directly addresses these gaps. We advance this agenda in three ways. First, and most critically, we analyze a unique dataset of 3,151 weekly voice diaries from 284 participants with major depression, allowing us to move beyond simple case-control classification and instead model the nuanced, longitudinal fluctuations of symptom severity as they occur in a real-world clinical context. Second, we bench-mark a multimodal feature set (classical lexical and basic prosodic metrics, TF-IDF, openSMILE acoustics, and Qwen3-8B sentence embeddings, a state-of-the-art 8-billion-parameter model) using a consistent support-vector regression protocol. Third, we pair the best-performing embeddings with BERTopic to expose the linguistic themes that drive prediction and to quantify their relationship with Beck Depression Inventory (BDI) scores.

Drawing on the ability of large language models to capture deep contextual information from text, we hypothesize that (i) Qwen3-8B embeddings will outperform all lexical and acoustic baselines in predicting continuous BDI scores. We further hypothesize that (ii) the resulting topic structure will reveal clinically coherent themes, contrasting expressions of internal distress with descriptions of external events, whose prevalence explains a substantial fraction of variance in concurrently measured symptom severity (BDI). By combining language representations from large-scale, billion-parameter models with unsupervised thematic analysis on a real-world corpus, our work aims to deliver a scalable and interpretable pipeline for continuous, voice-based depression monitoring.

## 2 Methods

### 2.1 Participants

The data for this study were drawn from a longitudinal project that used the ReMAP (Remote Monitoring Application in Psychiatry) smartphone application to acquire weekly speech samples from a cohort of both healthy controls (HC) and individuals with a lifetime or current diagnosis of Major Depressive Disorder (MDD). The full dataset consists of 3,151 weekly speech samples contributed by 284 unique participants (HC: 156; MDD: 128), collected between May 2019 and April 2025 over a mean period of 151 days per participant (median = 17 days, IQR = 0-137 days). On average, each participant provided 11.1 samples (SD = 28.5); however, the number of contributions was highly variable, with a median of 2 samples per participant (IQR = 1-7) and a maximum of 282 samples from a single individual. The study design, recruitment procedures, and the feasibility of digital data acquisition via the ReMAP smartphone application have been detailed in previous publications [17, 18].

Briefly, participants were recruited from ongoing mental health cohort studies. More than two-thirds of the participants in this dataset are part of the Marburg-Münster Affective Disorders Cohort Study (MACS) [19, 20], with data collected at two sites (Marburg and Münster, Germany) using identical study protocols. The MDD group comprised both acutely depressed individuals and those with a lifetime diagnosis who were in remission at the time of participation thus including a wide range of depression symptom severity. The presence or absence of a lifetime mental disorder was confirmed using the Structured Clinical Interview for DSM-IV (SCID-IV; [21]) as part of the original cohort protocols. The primary outcome measure for this analysis, depressive symptom severity, was the temporally closest Beck Depression Inventory (BDI) score collected within a ±7 day window of each speech sample. Demographic and clinical characteristics of the sample are presented in Table 1.

**Table 1.** Demographic and Clinical Characteristics of the Study Sample and Group Differences. MDD = Major Depressive Disorder; BDI = Beck Depression Inventory; SD = Standard Deviation. P-values from independent t-tests for continuous variables and Chi-squared tests for categorical variables.

| Characteristic | Healthy Controls | MDD | Total | Statistic |
| --- | --- | --- | --- | --- |
| No. of Participants,<br>N (%) | 156 (54.9%) | 128 (45.1%) | 284 (100.0%) |  |
| No. of Samples (n) | 1,880 | 1,271 | 3,151 |  |
| Age, years [Mean (SD)] | 53.04 (11.73) | 49.69 (11.45) | 51.70 (11.73) | $t = -7.31, p < .001$ |
| Sex, N (%) | | | | $\chi^2 = 65.28, p < .001$ |
| Female | 107 (37.7%) | 93 (32.7%) | 200 (70.4%) |  |
| Male | 49 (17.3%) | 35 (12.3%) | 84 (29.6%) |  |
| BDI Score at Time of<br>Sample [Mean (SD)] | 3.47 (4.29) | 12.14 (9.36) | 6.97 (8.03) | $t = -30.90, p < .001$ |

### 2.2 Data Acquisition

Data were collected using the ReMAP application, a native smartphone app developed for both iOS and Android platforms. The data acquisition protocol has been detailed in previous publications [18]. Following informed consent, participants installed the application on their personal smartphones, following a bring-your-own-device approach. The ReMAP app prompts users for active data collection, which includes weekly completion of the Beck Depression Inventory (BDI) and weekly voice samples. For the voice data collection central to this study, participants received randomized prompts during the day, asking them to speak freely for 1 to 3 minutes in response to the question: *“How did you feel during the last week?“* in German language.

### 2.3 Feature Extraction and Processing

To create a comprehensive feature set for modelling, we processed both the audio and the transcribed text from each speech sample.

#### 2.3.1 Speech-to-Text Transcription and Preprocessing

All audio samples were first transcribed locally using the open-source OpenAI Whisper (large-v2) model to generate verbatim transcripts [22]. From this raw text, a second, normalized version was created for specific downstream tasks. This cleaning process, performed using spaCy, involved converting text to lowercase, removing stop words and punctuation, and lemmatizing tokens.

#### 2.3.2 Acoustic Feature Extraction

From the raw audio files, we extracted two well-established, high-dimensional acoustic feature sets using the Python port of the openSMILE toolkit: the comprehensive ComParE functionals set (6,373 features; [23]) and the psychologically relevant eGeMAPS v02 set (88 features; [24]).

#### 2.3.3 Text-Based Feature Extraction

We extracted three distinct types of text-based features:

- **Lexical and Basic Prosodic Metrics:** A set of 18 features computed from the transcripts and audio timings, including metrics such as word count, lexical diversity, speech rate, and pause ratios. The complete list of lexical and prosodic metrics is provided in Table 2.
- **TF-IDF Features:** Term Frequency–Inverse Document Frequency vectors calculated from 1- and 2-word n-grams in the normalized transcripts; they weight each term by its rarity across the corpus, thus highlighting words and phrases that uniquely characterise an individual diary.
- **Sentence Embeddings:** State-of-the-art semantic representations were generated using the Qwen3-8B embeddings model. This open-source model was selected as its performance is comparable to leading proprietary models on public benchmarks (MTEB Leaderboard, checked 16 June 2025), while ensuring that sensitive participant data could be processed locally. To tailor the embeddings to our specific task, each verbatim transcript was prepended with the instruction: *“Generate a German embedding to analyse emotional and cognitive states from this weekly voice diary for a depression severity regression task.“* This ensures the model generates representations optimized for our goal while preserving the full contextual and grammatical structure of the speech.

**Table 2.** List of Lexical and Prosodic Metrics.

| Metric | Description |
| --- | --- |
| duration_seconds | Total duration of the audio recording. |
| speak_seconds | Total duration of speech segments. |
| word_count | Total number of words spoken. |
| char_length | Total number of characters in the transcript. |
| unique_word_count | Number of unique words used. |
| lexical_diversity | Ratio of unique words to total words. |
| avg_word_length | Average length of words. |
| words_per_total_second | Speech rate calculated over the total recording duration. |
| words_per_speak_second | Speech rate calculated over speech segments only. |
| speech_ratio | Proportion of the recording that is speech. |
| pause_seconds | Total duration of pauses. |
| pause_ratio | Proportion of the recording that is pauses. |
| pause_to_speech_ratio | Ratio of pause duration to speech duration. |
| entity_count | Number of named entities (e.g., persons, locations) identified. |
| entity_density | Ratio of named entities to total words. |
| noun_ratio | Proportion of words that are nouns. |
| verb_ratio | Proportion of words that are verbs. |
| adj_ratio | Proportion of words that are adjectives. |

### 2.4 Predictive Modelling

#### Predictive-modelling pipeline

We predicted the severity of depressive symptoms with a machine-learning pipeline that (1) standardised each feature matrix, (2) reduced dimensionality by principal-component analysis (PCA), and (3) fitted a support-vector regression (SVR) model. The ground-truth label was the Beck Depression Inventory (BDI) score recorded in the ReMAP app within *±* 7 days of the corresponding voice diary. A separate pipeline was trained for every single-modal feature set and for each predefined multimodal combination.

#### Software

All analyses were run in Python 3.11 using scikit-learn 1.6; PCA and SVR were GPU-accelerated with cuML 25.02.

#### Cross-validation and hyper-parameter tuning

Model performance was estimated with nested, participant-stratified five-fold cross-validation (GroupKFold in both outer and inner loops). The outer loop produced unbiased test estimates, whereas the inner loop tuned the SVR kernel, regularisation strength (C), *ɛ*-insensitive loss width, and the number of retained PCA components. Hyper-parameter search was orchestrated by Optuna’s median-pruner algorithm. Performance is reported as mean absolute error (MAE) and explained variance (*R*^2^).

#### Baselines

As points of reference we fitted (i) a DummyRegressor with strategy=“mean” for all regression tasks and (ii) a DummyClassifier with strategy=“stratified” for the HC vs. MDD supplementary classification.

#### Supplementary analyses

To contextualise the regression results we additionally (i) trained a binary classifier to distinguish healthy controls (HC) from participants with major depressive disorder (MDD), reporting balanced accuracy and AUROC, and (ii) repeated the full regression pipeline on the MDD subgroup alone to verify that performance reflected intra-group symptom variation rather than diagnostic differences.

#### Statistical significance

Significance was assessed with 1,000 label-shuffling permutations per feature set and 1,000 paired permutations comparing the leading models. Twotailed *p*-values were computed as (*r* + 1)*/*(*N* + 1), and all permutations were executed within the outer cross-validation folds to preserve the nested data-splitting structure.

### 2.5 Topic Modelling

To gain insight into the linguistic themes associated with depression severity, we conducted an unsupervised topic analysis using BERTopic [11] on the best-performing text embeddings (Qwen3-8B). The BERTopic pipeline employed UMAP (Uniform Manifold Approximation and Projection) for dimensionality reduction and HDBSCAN for clustering. The topic modelling was performed on the lemmatized transcripts to ensure coherent keyword representations. The resulting topics were manually labelled based on an inspection of each topic’s dominant keywords and a review of its most representative documents. After automatically reducing the number of topics to merge semantically similar clusters, we performed a series of statistical tests. A Kruskal-Wallis H-test was used to assess overall differences in BDI scores across topics, followed by pairwise Mann-Whitney U tests (with Benjamini-Hochberg FDR correction) to identify which specific themes differed significantly. Cliff’s Delta (*δ*) was calculated as a non-parametric measure of effect size.

## 3 Results

### 3.1 Predictive Modelling

We evaluated a range of feature modalities for their ability to predict continuous BDI scores (Table 3). All feature-based models substantially outperformed the dummy regressor (MAE = 6.24). The model based solely on Qwen3-8B sentence embeddings achieved an MAE of 4.45 and an *R*^2^ of 0.35, decisively outperforming every classical single-modality alternative (best classical: TF–IDF, MAE = 5.25, *R*^2^ = 0.19).

**Table 3.** Predictive Performance of Feature Sets for BDI Score Regression. All models are Support Vector Regressors with hyperparameters tuned via 5-fold nested cross-validation. MAE = Mean Absolute Error (lower is better); *R*^2^ = Coefficient of Determination (higher is better); SD = Standard Deviation across outer folds. Best single modality and multimodal combination are highlighted in bold.

| Feature Set | MAE (SD) | $R^2$ (SD) |
| --- | --- | --- |
| Any Feature Set (Dummy Baseline) | 6.24 (0.35) | -0.01 (0.05) |
| **Single Modality** |  |  |
| Lexical & Prosodic Metrics | 5.81 (1.04) | -0.15 (0.21) |
| TF-IDF | 5.25 (0.44) | 0.19 (0.07) |
| eGeMAPS acoustic | 5.61 (0.77) | -0.10 (0.13) |
| ComParE acoustic | 5.58 (0.74) | -0.04 (0.14) |
| **Qwen3-8B embeddings** | **4.45 (0.25)** | **0.35 (0.10)** |
| **Multimodal Combinations** |  |  |
| Lexical & Prosodic + TF-IDF | 5.37 (0.75) | -0.09 (0.14) |
| Lexical & Prosodic + eGeMAPS | 5.72 (1.02) | -0.14 (0.20) |
| Lexical & Prosodic + ComParE | 5.64 (0.89) | -0.10 (0.18) |
| TF-IDF + eGeMAPS | 5.74 (0.96) | -0.14 (0.19) |
| TF-IDF + ComParE | 6.24 (0.63) | -0.09 (0.11) |
| Qwen3-8B + Lexical & Prosodic | 4.42 (0.24) | 0.35 (0.11) |
| **Qwen3-8B + TF-IDF** | **4.39 (0.23)** | **0.35 (0.10)** |
| Qwen3-8B + eGeMAPS | 4.46 (0.27) | 0.35 (0.10) |
| Qwen3-8B + ComParE | 4.69 (0.33) | 0.29 (0.11) |

Adding shallow features yielded small but consistent gains. Two combinations, namely embeddings + lexical–prosodic metrics and embeddings + TF–IDF, both yielded modest performance improvements (best: MAE = 4.39, *R*^2^ = 0.35). Paired permutation tests confirmed that each of these multimodal models was significantly superior to embeddings alone (*p* = 0.021 and *p* = 0.018, respectively), although the absolute improvements were modest (ΔMAE < 0.07). No other fusion, acoustic or otherwise, matched this level of benefit.

Label-shuffling permutation tests (1,000 iterations per feature set) verified that every reported model performed above chance (*p <* 0.001). Effect sizes mirrored the descriptive results: embeddings delivered a large performance leap over the strongest classical baseline (ΔMAE = 0.80, Δ*R*^2^ = 0.16), whereas the multimodal additions provided only incremental refinement.

To benchmark the robustness of our findings, we conducted two validity checks. First, we trained binary classifiers to distinguish participants with Major Depressive Disorder (MDD) from healthy controls (HC). Results (see Table 4) showed that the best-performing models combined Qwen3-8B embeddings with either lexical–prosodic or eGeMAPS acoustic features, each achieving a balanced accuracy of 0.70. The embeddings alone reached 0.68, markedly surpassing the strongest classical baseline (TF–IDF, 0.62) and all acoustic-only models, confirming that the semantic information captured by large-scale embeddings carries a strong, readily extractable signal of depression status. Label-shuffling permutation tests confirmed that every classification model performed significantly above chance (*p <* 0.01), while a paired permutation test found no significant difference between the two leading multimodal models (balanced-accuracy Δ *≈* 0, *p >* 0.05).

**Table 4.** Classification performance (mean (SD) across outer folds) for distinguishing participants with major depressive disorder from healthy controls. Balanced accuracy and area under the ROC curve (AUC-ROC) are averaged across outer folds of nested cross-validation; the dummy baseline predicts the majority class.

| Feature Set | Balanced Accuracy (SD) | AUC-ROC (SD) |
| --- | --- | --- |
| Any Feature Set (*Dummy Baseline*) | 0.50 (0.00) | 0.50 (0.00) |
| Lexical & Prosodic Metrics | 0.59 (0.05) | 0.68 (0.04) |
| eGeMAPS acoustic | 0.55 (0.03) | 0.55 (0.08) |
| ComParE acoustic | 0.59 (0.05) | 0.60 (0.14) |
| TF-IDF | 0.62 (0.03) | 0.70 (0.07) |
| Lexical & Prosodic + TF-IDF | 0.62 (0.04) | 0.70 (0.04) |
| Qwen3-8B Embeddings | 0.68 (0.06) | 0.76 (0.06) |
| **Qwen3-8B + Lexical & Prosodic** | **0.70 (0.03)** | **0.77 (0.04)** |
| Qwen3-8B + eGeMAPS acoustic | 0.70 (0.04) | 0.77 (0.05) |

Second, to test whether our models were simply separating healthy controls from patients rather than tracking symptom variation within a clinical cohort, we repeated the entire predictive-modelling pipeline using only the 1 271 diaries contributed by the 128 MDD participants (Table 5). In this more challenging setting the Qwen3-8B embeddings were the only feature set to demonstrate predictive power, achieving an MAE of 6.01 and an *R*^2^ of 0.15. In contrast, all other feature modalities (lexical, acoustic, and TF-IDF) performed at, or slightly below, the dummy baseline (MAE = 7.37; *R*^2^ = –0.06).

**Table 5.**
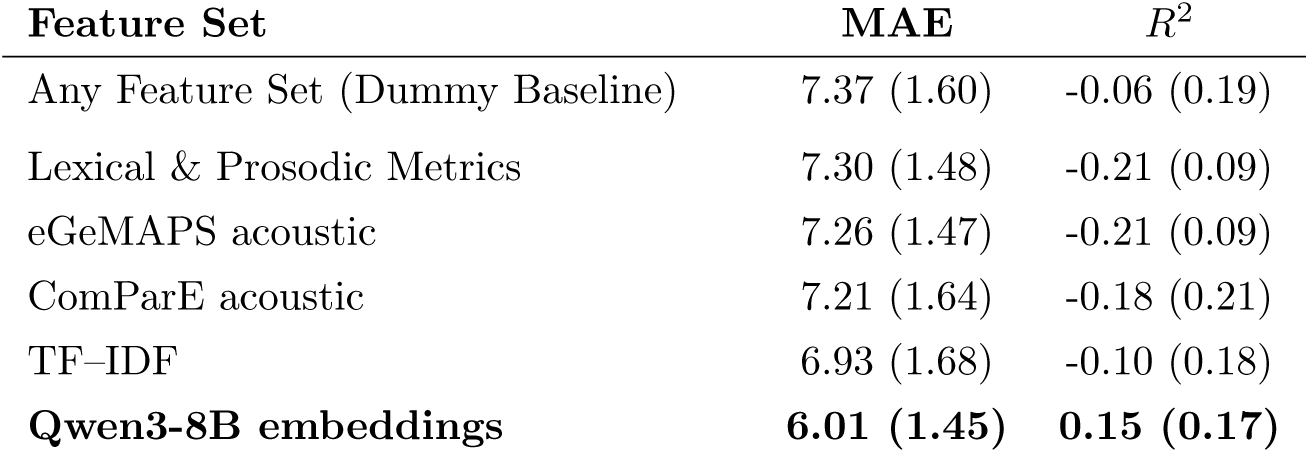
Predictive Performance within MDD Cohort Only (N=128). All models are Support Vector Regressors with hyperparameters tuned via 5-fold nested cross-validation. MAE = Mean Absolute Error; *R*^2^ = Coefficient of Determination. Best performance is highlighted in bold.

### 3.2 Topic Modelling

To understand the linguistic content driving the predictive performance, we conducted an exploratory topic analysis using BERTopic. This data-driven approach yielded 10 distinct, interpretable topics within the speech diaries that we labelled according to the dominant keywords and most representative documents (see Table 6). A Kruskal-Wallis H-test revealed a highly significant association between BDI scores and these discovered topics (H = 663.92, p < .001).

**Table 6.** Topic Summary and BDI Statistics. Topics are ordered from highest to lowest Mean BDI score. Documents (n) = number of voice diary entries assigned to each topic; Subjects (N) = number of unique participants contributing to each topic. The outlier topic has been excluded from this analysis.

| Topic Name | Rationale (based on key words & representative documents) | Documents (n) | Subjects (N) | Mean BDI (SD) |
| --- | --- | --- | --- | --- |
| Persistent Low Mood | Mentions of chronic tiredness and ongoing therapy indicate a lingering dysphoric state conveyed with hesitant phrases like *halt* and *irgendwie*. | 311 | 19 | 14.03 (7.40) |
| Pain Distress | *Schmerz* dominates, alongside talk of operations, health worries, and exhaustion, highlighting acute somatic discomfort driving emotional strain. | 45 | 9 | 12.24 (4.55) |
| Routine Stress | Descriptions of ordinary work weeks, brief holidays, and household chores reflect manageable but recurring everyday pressures. | 880 | 203 | 6.61 (6.95) |
| Rebuilding Stability | Narratives about moving, setting up a new home, and regaining strength (*Kraft aufbauen*, *stabil*) portray a constructive recovery phase. | 46 | 3 | 3.98 (5.55) |
| Heavy Workload | Focus on budget planning, restructuring, and *Kurzarbeit* shows high task load and organizational change rather than emotional content. | 57 | 4 | 3.75 (2.06) |
| Social Joy | Stories of birthdays, vacations, good weather, and family time emphasize pleasant social and seasonal experiences. | 67 | 22 | 3.37 (4.59) |
| Casual Updates | Conversational check-ins with minor ailments (e.g., mild COVID colds) and everyday remarks resemble informal status updates. | 47 | 7 | 2.91 (6.60) |
| Travel & Partner | Recurrent trips to Würzburg and time with a partner center on logistics and companionship in daily routine. | 144 | 7 | 2.88 (0.80) |
| Rehab Progress | Detailed accounts of shoulder/knee therapy, orthoses, and physiotherapy track physical rehabilitation milestones. | 67 | 9 | 1.51 (2.00) |
| Planning & Leisure | Entries about lesson prep, timetables, weekend walks, and family dinners highlight structured planning balanced with enjoyable downtime. | 153 | 5 | 0.67 (1.92) |

Post-hoc pairwise comparisons (Mann-Whitney U tests with FDR correction) confirmed that specific topics were strongly linked to symptom severity. In particular, themes reflecting emotional distress such as “Persistent Low Mood” (Mean BDI = 14.03) and “Pain Distress” (Mean BDI = 12.24) were associated with the highest depression scores. The most pronounced difference was found between “Persistent Low Mood” and the activity-focused topic “Planning & Leisure” (Mean BDI = 0.67), which yielded the largest effect size in the analysis (Cliff’s *δ* = 0.99, p < .001). The distribution of BDI scores for each topic is visualized in Figure 1. To visualize the relationships between these themes, we computed their pairwise semantic similarity, revealing the distinct topical structure of the diaries (Figure 2).

**Figure 1.**
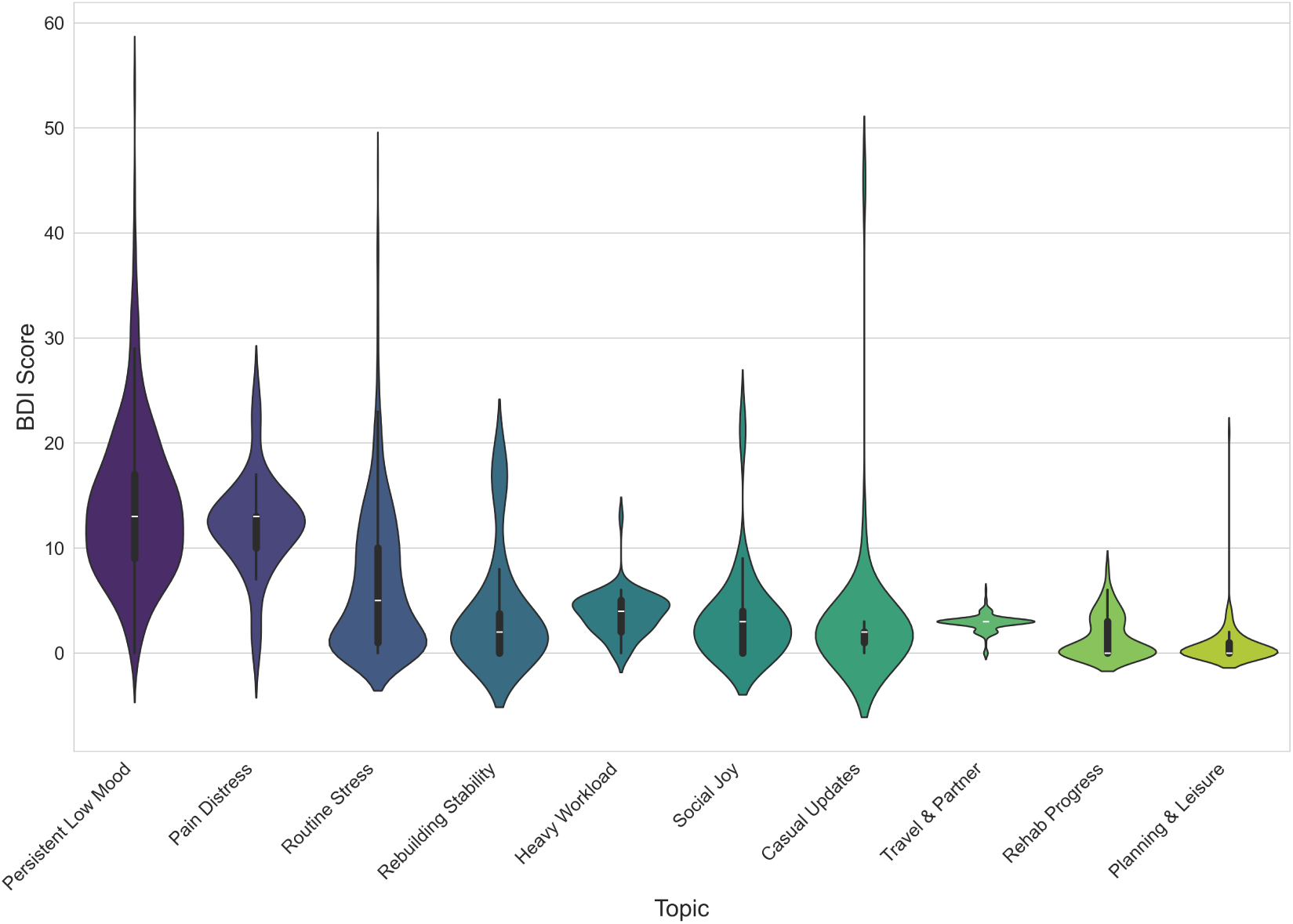
Distribution of Beck Depression Inventory (BDI) scores across 10 machine-learning-derived topics. The topics are arranged as violin plots along the horizontal axis in descending order of their mean BDI score. This ordering reveals a clear severity gradient. The leftmost topics, such as “Persistent Low Mood” and “Pain Distress”, are visibly associated with higher BDI scores, whereas the rightmost topics, centered on daily activities like “Planning & Leisure”, correspond to lower scores.

**Figure 2.**
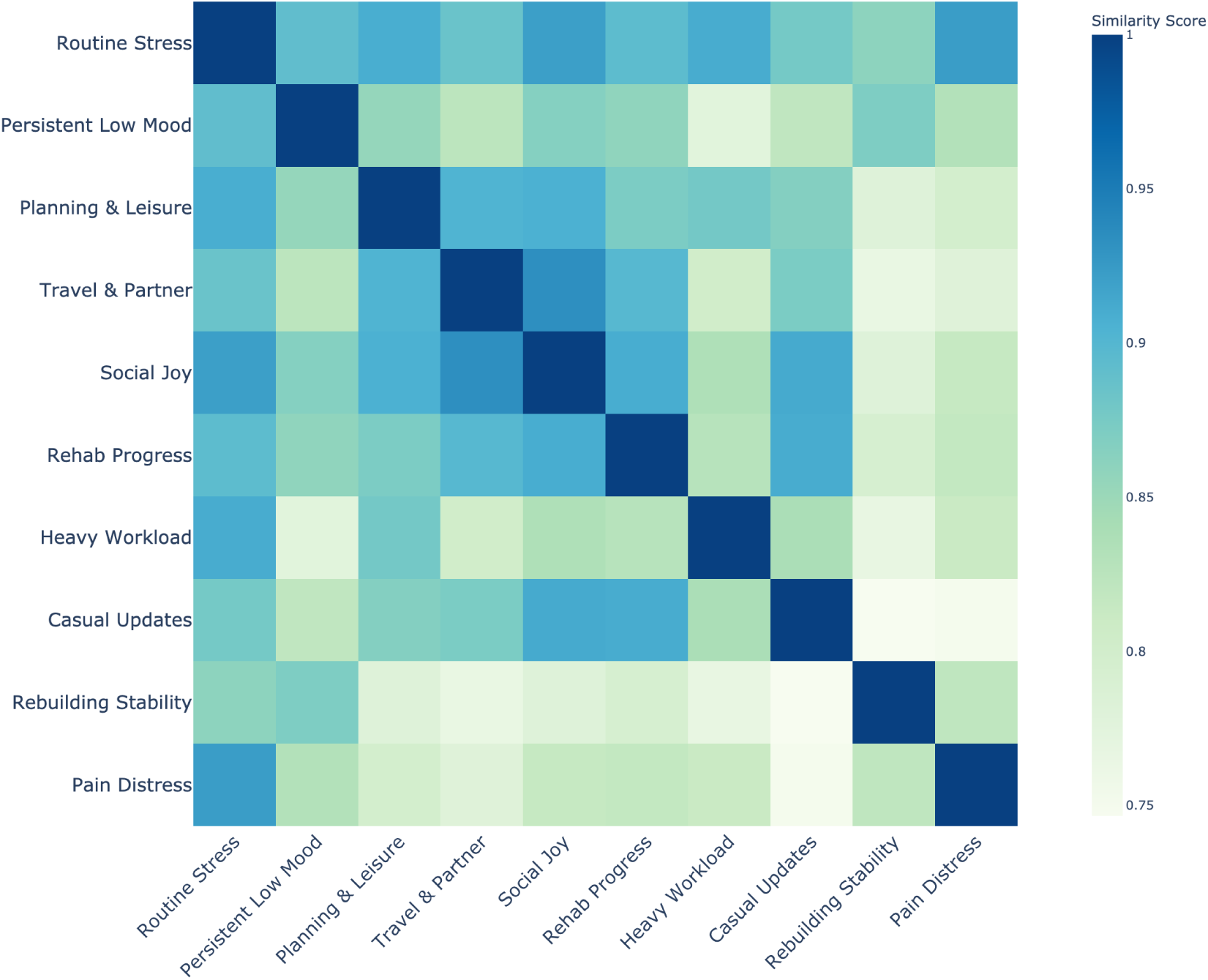
Pairwise cosine similarities among the ten BERTopic themes are displayed as a hierarchically ordered heat-map (Qwen3-8B embeddings; diagonal fixed at 1.0). A compact high-similarity block (deep blue) links the routine narratives Routine Stress, Social Joy and Casual Updates, while a second, looser block groups the domestic-routine themes Planning & Leisure and Travel & Partner. In contrast, the health-related topics diverge markedly: Rehab Progress shows only moderate affinity to Pain Distress, and Rebuilding Stability occupies the most isolated position in the matrix, underscoring its distinct lexical profile. The pale band separating these regions confirms the semantic distance between day-to-day event descriptions and more introspective health reflections, complementing the super-cluster pattern observed in the UMAP projection.

To quantify the predictive utility of these themes alone, we trained a separate regression model using only the topic probabilities as features. This topic-only model was able to predict BDI scores with an MAE of 5.68 ± 0.77 on 5-fold cross-validation, demonstrating that the discovered topics themselves capture a substantial and clinically meaningful portion of the variance in depression severity.

## 4 Discussion

This study developed and evaluated a pipeline for predicting depressive symptom severity from repeated, naturalistic voice diaries. Our findings yielded two principal insights: first, that large-scale sentence embeddings significantly outperform classical text and acoustic features for this task; and second, that an exploratory topic analysis of these embeddings can reveal clinically coherent themes that are strongly associated with depression severity.

Our primary predictive finding is the clear superiority of the Qwen3-8B sentence embeddings over all classical feature approaches. This result strongly supports the growing consensus that the semantic content of speech, as captured by large language models, is the dominant signal for detecting depressive states. While adding either lexical–prosodic or TF–IDF features yielded slightly better scores (best: embeddings + TF–IDF, MAE = 4.39; *R*^2^ = 0.35), this represents only a modest improvement, suggesting that embeddings capture the vast majority of the predictive signal. The small additional contribution from features like speech duration and pause ratios warrants further investigation, but it may indicate that basic speech timing and word rarity provide complementary, non-semantic information. Our findings clearly demonstrate that modern language model representations have fundamentally shifted the landscape of speech-based depression detection. Our sensitivity analysis, which evaluated the model exclusively on the MDD cohort, further strengthens this conclusion. This result strongly suggests that while many features can distinguish between healthy and depressed states, only the rich semantic representations from the Qwen3-8B embeddings are capable of capturing the nuanced variance in symptom severity within a clinical population.

A central contribution of this study is to show that high-performance models can be made interpretable rather than remaining inscrutable “black boxes”. By applying BERTopic to the embeddings, we moved beyond prediction and towards interpretation. Our data-driven approach independently uncovered topics that align closely with established clinical knowledge. The strong association between higher BDI scores and themes like “Pain Distress” and “Persistent Low Mood” suggests our model is sensitive to both somatic complaints and the language of internal coping or distress. This finding corroborates previous work showing that themes of self-focus and negative cognitive patterns are robust linguistic markers of depression [25–27].

Crucially, the ability of a separate model to predict BDI scores using only these topic probabilities confirms their clinical relevance. This dual approach of using embeddings for prediction and topic modelling for interpretation offers a powerful model for future research. It allows for the development of accurate screening tools while also providing a pathway for clinicians to understand the potential drivers of a high predicted score, thereby fostering trust and aiding therapeutic dialogue [28].

The present findings highlight the potential of using speech samples collected in naturalistic settings via participants’ personal devices, combined with advanced natural language processing techniques, for monitoring depressive symptoms. This approach offers a scalable and ecologically valid method for digital phenotyping in mental health. Future research should investigate the generalizability of these findings across diverse populations, contexts, and technological platforms. Given the potential utility of such tools, further work is warranted that includes patient and public involvement and integrates the perspectives of caregivers and clinicians. This would help identify feasible and acceptable pathways for implementing speech-based monitoring in both research and clinical practice. Although not directly assessed in this study, it is worth considering the possibility that speech-based assessments may be perceived as less burdensome than traditional methods, such as text-based patient-reported outcomes or ecological momentary assessments. The act of recording speech can be conveniently integrated into daily activities, a concept already applied in emerging clinical decision support systems such as DAX Copilot^1^, which could potentially improve user engagement and adherence.

Several limitations should be acknowledged. First, our study relies on self-reported BDI scores as the primary outcome measure. While we previously demonstrated high correspondence between smartphone-derived self-reports via ReMAP and external clinical ratings of depression severity [17], the reliance on self-report remains a methodological consideration. Second, our findings are based on a German-speaking cohort recruited primarily from two metropolitan areas (Marburg and Münster) using a bespoke research application (ReMAP). The specific demographics and clinical characteristics of this sample may therefore limit the broader generalizability of our findings to other languages, cultures, and healthcare systems. Third, our analysis is fundamentally correlational in nature; while we can demonstrate strong associations between linguistic themes and BDI scores, we cannot establish causal relationships between specific language patterns and depressive symptoms. Finally, the naturalistic data collection approach, while ecologically valid, introduces variability in recording conditions and speech content that may affect model performance in more controlled clinical settings.

This study demonstrates that modern, open-source language models can predict depression severity from repeated, smartphone-collected voice diaries. We have shown that this predictive power is not opaque; it is driven by interpretable, clinically relevant linguistic themes that can be discovered directly from the data. By combining state-of-theart language model embeddings with data-driven interpretation, this work represents a meaningful step towards developing scalable, non-invasive, and understandable tools for monitoring mental health in the real world.

## Data Availability

As data was derived from multiple study protocols with different limitations, the potential sharing of data would need to be discussed and evaluated on a case-by-case basis with the senior authors and different study leaders

## Acknowledgements

This work was funded in part by the consortia grants from the German Research Foundation (DFG) FOR 2107 and SFB/TRR 393 (project grant no 521379614), as well as the DYNAMIC center, funded by the LOEWE program of the Hessian Ministry of Science and Arts (grant number: LOEWE1/16/519/03/09.001(0009)/98).

## Author Contributions

TH and NO had the initial idea of the ReMAP app and designed the project from which these data are drawn. DE conceived and designed the study, developed the ReMAP app, implemented all preprocessing and modelling code, performed statistical analyses, created visualisations and wrote the main manuscript text. MR, AC, RL, JH, VH and EL contributed to study design, data interpretation and substantive manuscript revisions. LG, AF, RB, JG, NW, JS, SM, TB, KF, FS, NA and HJ coordinated participant recruitment, collected speech diaries and associated clinical data, performed data quality assurance and edited the manuscript. JR, CD, RR, UEP, IN, TK, UD, TH and NO provided clinical oversight, project supervision and edited the manuscript. All authors provided critical feedback and approved the final version. DE and NO are the guarantors of the study.

## Footnotes

1 Microsoft Dragon Copilot | Microsoft Cloud for Healthcare. Available at: https://www.microsoft.com/en-us/health-solutions/clinical-workflow/dragon-copilot

## Notes

### Competing Interest Statement

The authors have declared no competing interest.

### Author Declarations

Ethik-Kommission der Aerztekammer Westfalen-Lippe und der Westfaelischen Wilhelms-Universitaet, Muenster, Germany gave ethical approval for this work.

### Summary of Updates

Title shortened to remove "non-invasive" for concision; author list expanded (adds Jonathan Repple and Christian Dobel) and multiple affiliations corrected; Abstract rewritten with updated metrics (MAE = 4.45, R2 = 0.35) and new comparison highlighting a 16-point gain over TF-IDF instead of the earlier "modest improvement over embeddings" wording; Methods section now specifies software versions, Optuna search space and 1,000-iteration permutation tests; Results section integrates binary HC-vs-MDD classification (Table 4) and within-MDD sensitivity analysis (Table 5) that were previously in the Supplement; wording throughout adjusted for clarity, consistent terminology; all tables and figures renumbered and Figure 2 updated (heatmap now in main text); supplementary material removed.

